# Arbovirus risk perception as a predictor of mosquito-bite preventive behaviors in Ponce, Puerto Rico

**DOI:** 10.1101/2022.04.27.22274353

**Authors:** Josée M. Dussault, Gabriela Paz-Bailey, Liliana Sánchez-González, Laura E. Adams, Dania M. Rodríguez, Kyle R. Ryff, Chelsea G. Major, Olga Lorenzi, Vanessa Rivera-Amill

## Abstract

Mosquito-borne arboviruses are an important cause of morbidity and mortality in the Caribbean. In Puerto Rico, chikungunya, dengue, and Zika viruses have each caused large outbreaks during 2010–2022. To date, the majority of control measures to prevent these diseases focus on mosquito control and many require community participation. In 2018, the U.S. Centers for Disease Control and Prevention launched the COPA project, a community-based cohort study in Ponce, Puerto Rico, to measure the impact of novel vector control interventions in reducing arboviral infections. Randomly selected households from 38 designated cluster areas were offered participation, and baseline data were collected from 2,353 households between May 2018 and May 2019. Household-level responses were provided by one representative per home. Cross-sectional analyses of baseline data were conducted to estimate 1) the association between arboviral risk perception and annual household expenditure on mosquito control, and 2) the association between arboviral risk perception and engagement in ≥3 household-level risk reduction behaviors. In this study, 27% of household representatives believed their household was at high risk of arboviruses and 36% of households engaged in at least three of the six household-level preventive behaviors. Households where the representative perceived their household at high risk spent an average of $35.9 (95% confidence interval: $23.7, $48.1) more annually on mosquito bite prevention compared to households where the representative perceived no risk. The probability of engaging in ≥3 household-level mosquito-preventive behaviors was 10.2 percentage points greater (7.2, 13.0) in households where the representatives perceived high risk compared to those in which the representatives perceived no risk. Paired with other research, these results support investment in community-based participatory approaches to mosquito control and providing accessible information for communities to accurately interpret their risk.

**Author Summary:** Mosquito-borne disease is an important cause of illness and death in the Caribbean, including Puerto Rico. Most tactics to prevent these diseases rely on stopping mosquito bites, either by reducing the mosquito population or creating barriers between mosquitos and humans. These methods vary in the degree of community involvement required. This study used data collected from 2,353 households in Ponce, Puerto Rico from May 2018 to May 2019 to understand how household perception of risk of contracting these diseases related to 1) the amount of money households spent annually to prevent mosquito bites, and 2) the number of activities their household engaged in to prevent mosquito bites. We found that 27% of households perceived themselves at high risk of contracting these diseases, and 36% of households engaged in at least three activities to reduce their risk. On average, households that perceived themselves at high risk spent more money on mosquito bite prevention and engaged in more activities to prevent mosquito bites, compared to households that perceived no risk. Paired with other research in this area, these results support investment in community-based approaches to mosquito control and ensuring that communities have accessible information to understand their risk of mosquito-borne disease.

## Introduction

Mosquito-borne illnesses are an important cause of morbidity and mortality in the Caribbean [1]. In Puerto Rico, chikungunya, dengue, and Zika viruses have each caused large outbreaks during 2010–2022. Although chikungunya and Zika viruses were recently introduced to Puerto Rico, dengue is endemic and seasonal, with outbreaks occurring every 3–5 years [1,2]. All three viruses are transmitted via *Aedes* species mosquito vectors and can cause acute febrile illness, although each of these diseases has the potential for severe or long-term outcomes [2–4]. Patients with chikungunya can have persistent joint pain for months or years. Perinatal Zika infections are known to cause birth defects [5,6]. Evidence suggests that individuals infected with chikungunya or Zika virus are conferred immunity against the respective virus [5,7]. However, dengue virus has four serotypes, and individuals who have been infected with one serotype have a higher risk of severe dengue if later infected with a different serotype. Severe dengue can be lethal and is characterized by severe plasma leakage, severe hemorrhage, and/or organ impairment [8]. There are no specific treatments for any of these diseases and management is primarily supportive.

The lack of treatment options for arboviruses is compounded by the scarcity of pharmaceutical prevention methods. There is currently one vaccine for dengue, CYD-TDV, approved by the U.S. Food and Drug Administration (FDA) and recommended by the Advisory Committee on Immunization Practices (ACIP) for use in children 9–16 years old who live in an endemic area and who have a lab-confirmed history of a previous dengue infection [9,10]. However, CYD-TDV is only available to a small proportion of the population at risk for dengue [10], and there are no other FDA-approved vaccines to protect people from these arboviruses. Thus, the public health community relies on vector control tools to prevent or reduce arboviral infections in endemic populations. Some of these vector control tools focus on killing adult mosquitos or reducing population growth through methods like disposal of stagnant water to decrease *Aedes* mosquito breeding sites, fumigation, and mosquito traps. Other methods focus on creating barriers between humans and mosquitoes through the use of screens in the doors and windows of buildings, centralized air conditioning, topical mosquito repellents applied to skin or clothing, and bed nets.

There are several barriers to mosquito-borne disease prevention, both at the individual and household level. In Puerto Rico, where the median household income was $20,296 in 2018 [11], the cost of supplies and services to reduce mosquito populations and mosquito bites can represent a considerable financial burden. Moreover, individuals may perceive themselves at low risk of contracting arboviral disease, either because they do not see illnesses in their communities or do not feel that mosquitos are a problem [12]. People who perceive themselves at low risk may be less motivated to protect themselves and other members of their household from contracting an arboviral infection. Whether due to financial capacity or motivation, the underutilization of mosquito prevention techniques and services puts populations at higher risk of arboviral infection.

Most models of health behavior generally emphasize the role of risk perception in decision making [13,14], but empirical studies on arboviral risk perception have produced conflicting results [12,15], and there have been few studies investigating arboviral risk perception and health behaviors in Puerto Rico [16]. Understanding the association between risk perception and risk reduction behaviors among people living in Puerto Rico can help guide selection of appropriate public health messaging and interventions to reduce the incidence of arboviral diseases. If risk perception correlates to increased protective behaviors, increasing campaigns to raise community awareness about arboviral disease risks could be beneficial. Alternatively, in the presence of a null relationship between risk perception and engagement in protective behaviors, then public health agencies may find it more effective to focus efforts on disease prevention methods that do not require active community engagement.

Accordingly, the objective of this study is to better understand the relationship between risk perception of arboviral diseases and adoption of behaviors to prevent mosquito bites among people living in southern Puerto Rico communities with high historical arboviral disease incidence rates. The central hypothesis is that individuals who perceive themselves at high risk for arboviral infection will be more likely to engage in mosquito-preventive behaviors than those who perceive no risk. Specifically, this study aims to (1) estimate the association between perceived household risk of arboviral infection and annual household expenditure on mosquito control, and (2) estimate the association between perceived household risk of arboviral infection and reported engagement in mosquito-controlling behavior (i.e., spraying insecticide, burning citronella candles/coils, etc.) among households in Ponce, Puerto Rico.

## Methods

### Parent Study

COPA (Communities Organized to Prevent Arboviruses) is a prospective community-based cohort study in Ponce, Puerto Rico initiated in 2018 to evaluate the impact of novel vector control interventions in reducing arboviral infections. Households were selected randomly from 38 study-defined cluster areas and visited up to three times to invite them to participate in COPA; field-level strategies are detailed elsewhere [17]. From May 2018 to May 2019, 2,353 households were enrolled in COPA. Household residents were eligible if they slept an average of four or more nights per week in the home, were 1-50 years old, and did not intend to move in the next six months. Study staff administered a standardized questionnaire to participants in Spanish regarding their knowledge, attitudes, and practices relating to arboviral infections and prevention methods. For each household, we asked one resident (an adult or emancipated minor) present during data collection to act as the household representative and answer an additional series of questions specific to their household. The following is an analysis of cross-sectional interview data from the COPA household representatives.

### Exposure

The exposure of interest was the household representative’s perception of their household’s risk of contracting arboviral illnesses in the next 12 months, which was assessed separately for Zika, dengue, and chikungunya. However, as the responses to these questions were highly correlated, dengue was used as a surrogate for all three arboviruses to measure the household representatives’ perceived risk of arboviral infection with three levels: high risk, low risk, and no risk. Models were tested in which the exposure was coded as a single 3-level ordinal variable versus as a set of two disjoint indicator (dummy) variables. Disjoint indicator variables allowed the model to relax the assumption of linearity between the three levels of the exposure variable by creating two binary variables that represented the low risk and high risk groups, with no risk as the referent level. The Bayesian information criterion (BIC) and Akaike information criterion (AIC) were used to compare the fit of the two models. Generally, models with lower AIC and BIC values are preferred. Where the values produced different conclusions, the BIC was preferred rather than the AIC because it usually selects the correct model more frequently than the AIC [18].

### Outcomes

There were two outcomes of interest. The first outcome of interest was annual household expenditure on mosquito prevention products and services, reported in United States dollars (USD) by the household representative. The second outcome was regular engagement in three or more mosquito preventive behaviors. In total, there were six possible household-level behaviors that the household representative could report during the baseline interview: (1) Eliminating stagnant water around the house (ex: cleaning containers that collect water like flowerpots and/or gutters; (2) Covering containers that collect water; (3) Emptying trash cans, drums, or in-ground trash cans if they had standing water; (4) Cleaning or removing tires and/or debris from the yard; (5) Spraying insecticide or fumigating indoors or outdoors; and (6) Burning citronella candles and/or mosquito coils. These behaviors were reported as the frequency with which any members of the household engaged in these activities in the previous 12 months, with responses including “daily”, “weekly”, “monthly”, “rarely”, or “never”. To dichotomize each of the six activities, households that reported daily or weekly engagement in these behaviors were counted as engaging in these behaviors regularly, and those who reported performing the activities monthly, rarely, or never were categorized as not conducting these activities regularly. This outcome was then dichotomized as regular engagement in three or more of these behaviors. The cutoff of three behaviors was selected because some activities (e.g., behaviors 1 and 3) appeared to be correlated, likely because it would be easy to accomplish one task while completing the other.

### Covariates

Covariates included in this study were from the household representatives’ responses to questionnaires from the same study visit. These variables included: mosquito bite frequency, household income, exposure to community educational campaigns, education level, sex, and the community cluster to which the household belonged. Reported mosquito bites was a binary variable produced from a COPA question asking about where the participants most often were bitten. If the participant responded saying that they rarely/never were bitten, this was encoded into the bite frequency variable as “never bitten”. Otherwise, participants were categorized as “bitten”. Household income was reported as one of eight categories and remained an ordinal categorical variable in statistical analyses. Education level was collapsed into five categories due to small counts in some of the original categories. Sex and educational campaign exposure were binary variables, and age was maintained as a continuous variable. Education level and community cluster were modeled as disjoint indicator variables. Rooted in Health Belief Model theory and additional readings [12–16,18–28], we developed directed acyclic graphs (DAGs) that assisted in the identification of mediating paths and confounders [29–32]. Based on these DAGs, the aforementioned covariates included in this study represented a sufficient set of confounders. Because participating households were selected from community clusters, the intra-cluster correlation (ICC) coefficient was calculated to assess the need to use multilevel modeling techniques; the ICC was determined to be small enough (0.02) to include community clusters as disjoint indicator variables rather than using a 2-level model [33].

### Missing Data

Approximately 11% of participants were missing data on at least one of the covariates, and most missing data came from the household income variable. A complete case analysis of these data could result in loss of precision and validity by not including the full possible sample of households [34–39]. Thus, this analysis used multiple stochastic conditional-mean imputation with coefficient resampling to assign values onto all missing covariates based on each participant’s observed characteristics, assuming that data were missing at random, conditional on the additional covariates in the imputation model [37,38,40]. The covariates included as predictive variables in the imputation models were employment status, whether the house had screens on the windows, and the six risk reduction behaviors described in the outcomes section. All exposure, outcome, and confounding variables that would be included in the analysis model were also included in the multiple imputation model. Multiple imputation methods were applied using Markov chain Monte Carlo (MCMC) method in SAS software, version 9.4.9 [41], with 50 imputed datasets, 500 burn-in iterations, and 500 additional iterations between imputations; the method used a single chain. The MCMC method was necessary because the missing data structure was arbitrary, and the number of burn-in iterations, additional iterations, and imputed datasets were selected based on recommendations to reduce Monte Carlo error [42–44]. Because two functional forms for the exposure variable were compared using model fit statistics that are not available in multiple imputation models, complete case models were used to assess the proper functional form of the exposure variable, and the multiple imputation model with that exposure coding was preferred over the complete case model.

### Analytical approach

#### 1. Household expenditure

In this analysis, we were interested in understanding the association between arboviral disease risk perception and annual household expenditure on risk reduction products and services as the primary outcome. To estimate the association, we fit a normal linear regression model. To meet the assumption of homoscedasticity in the model residuals [45], five outlier households whose expenditures ranged from $1800 to $3600 were set to missing values. For comparison, the sixth-largest expenditure value in the data was $1440, and the mean expenditure amount was $185. In the regression model, we included the following potential confounders: reported mosquito bites, household income, exposure to community educational campaigns, sex and education level of the household representative, and the community cluster to which the household belonged.

#### 2. Household-level risk reduction

The objective of this analysis was to understand the association between household perception of arboviral disease risk and the prevalence of engaging in household-level risk reduction behaviors. To estimate the prevalence difference, we fit a linear binomial model including the same set of potential confounders as described previously. Missing data on all variables were addressed using multiple imputation as described above. Data were analyzed using SAS software, version 9.4.9 [41].

### Ethical Consideration

All participants provided written, informed consent. The COPA study was approved by the Institutional Review Boards (IRB) of Ponce Medical School Foundation, Inc., and CDC. An exemption for this secondary statistical analysis was approved by the IRB at the University of North Carolina at Chapel Hill.

## Results

Among 2,353 household representatives who provided responses, 635 (27%) perceived their household at high risk of contracting dengue in the next 12 months. Comparatively, 1,249 (53%) believed their households were at low risk, and 425 (18%) perceived no risk (Table 1). The mean age of household representatives was 37 years (standard deviation [SD]: 8.8 years), 1,596 (68%) were female, and 1,429 (61%) reported having completed some postsecondary schooling. Most household representatives reported that they were bitten by mosquitoes (n=2,304; 98%); few reported that they were rarely or not bitten (n=45; 2%). The most commonly reported household annual income category was <$10,000 (n=915; 39%). Mean annual expenditure on mosquito products was $185 (SD: $225). A majority (n=1,884; 80%) of household representatives perceived at least some risk of contracting dengue in the next year (Tables 1 & 2).

**TABLE 1.** Distribution of COPA Household Representatives’ Perceived Risk by Arbovirus (N=2,353), 2018-2019.

|  | Dengue, N (%) | Zika, N (%) | Chikungunya, N (%) |
| --- | --- | --- | --- |
| **Perceived high risk** | 635 (28) | 606 (26) | 613 (27) |
| **Perceived low risk** | 1249 (54) | 1264 (55) | 1221 (53) |
| **Perceived no risk** | 425 (18) | 435 (19) | 473 (21) |
| **Missing** | 44 | 48 | 46 |
| **Total** | 2309 | 2305 | 2307 |

Household representatives who perceived themselves at no risk of arboviral diseases generally had lower levels of education compared to the low-risk and high-risk perception groups. Contrary to expectation, the no-risk perception group was also more likely to report having seen an educational campaign on mosquito prevention and control in the last 12 months compared to the other two risk perception groups. In total, 1,318 (56%) household representatives reported using mosquito repellent in the last 30 days (Table 2).

**TABLE 2.**
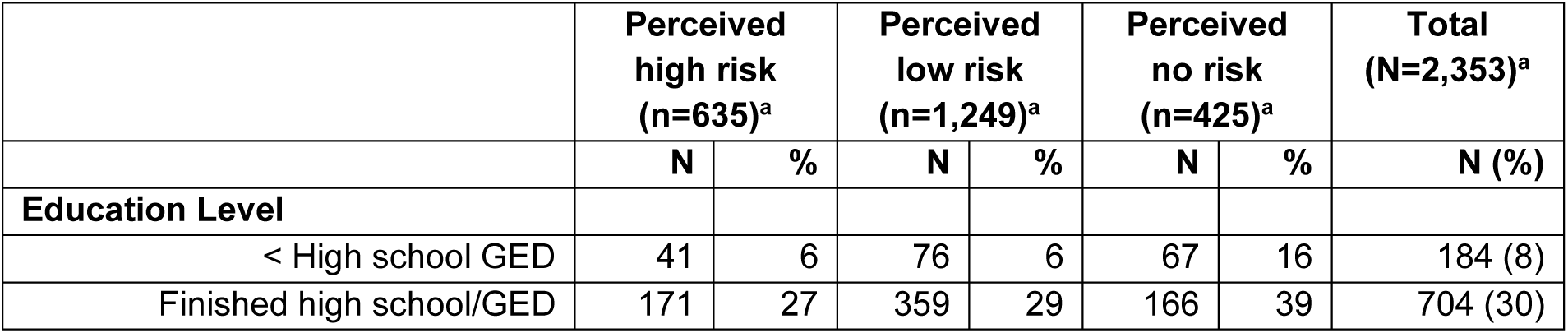
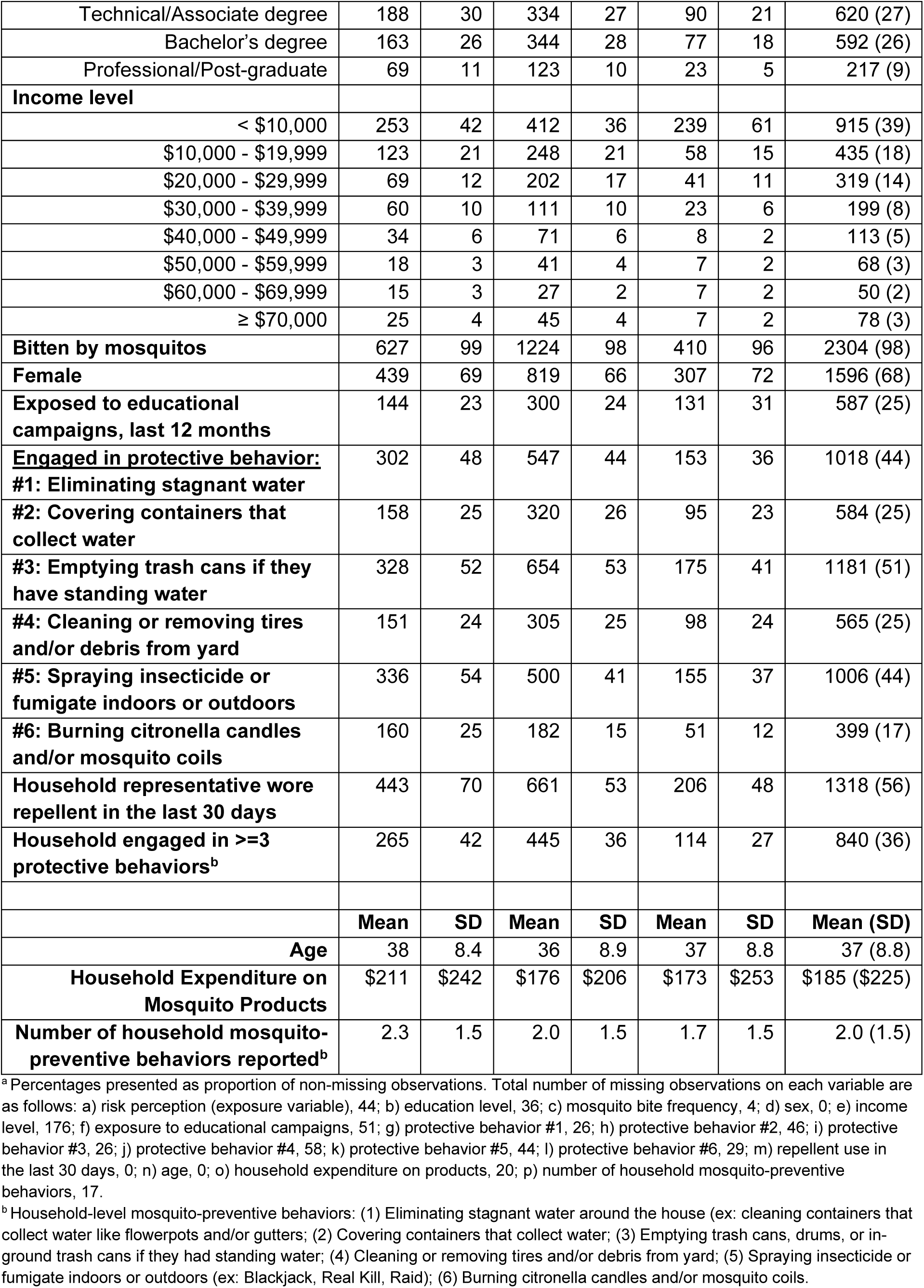
Characteristics of COPA household representatives by perceived risk of dengue (N=2,353), 2018-2019.

There was generally low uptake of the six household-level preventive behaviors assessed. About half of household representatives (51%) reported emptying trash cans with standing water, 44% reported eliminating other sources of stagnant water, and 44% reported spraying insecticide around the house or fumigating indoors or outdoors. A small number of households reported covering containers that could collect water (25%), cleaning debris from around the outside of the house (25%) or burning citronella candles or coils (17%) (Table 2). In total, 840 (36%) households engaged in at least three of the six household-level preventive behaviors. Households that were engaged in three or more preventive behaviors generally had household representatives with higher levels of education, a larger household income, and spent more money on mosquito products compared to households that engaged in fewer than three preventive behaviors (Table 3).

**TABLE 3.** Characteristics of COPA household representatives by protective behavior level (N=2,353), 2018-2019^a^.

|  | Engaged in < 3 protective behaviors (n=1,496)<sup>b</sup> |  | Engaged in ≥ 3 protective behaviors (n=840)<sup>b</sup> |  | Total (N=2,353)<sup>b</sup> |
| --- | --- | --- | --- | --- | --- |
|  | N | % | N | % | N (%) |
| **Education Level** |  |  |  |  |  |
| < High school GED | 134 | 9 | 49 | 6 | 184 (8) |
| Finished high school/GED | 485 | 33 | 215 | 26 | 704 (30) |
| Technical/Associate degree | 370 | 25 | 245 | 30 | 620 (27) |
| Bachelor's degree | 353 | 24 | 235 | 28 | 592 (26) |
| Professional/Post-graduate | 134 | 9 | 83 | 10 | 217 (9) |
| **Income level** |  |  |  |  |  |
| < \$10,000 | 634 | 42 | 281 | 33 | 915 (39) |
| \$10,000 - \$19,999 | 251 | 17 | 184 | 22 | 435 (18) |
| \$20,000 - \$29,999 | 198 | 13 | 121 | 14 | 319 (14) |
| \$30,000 - \$39,999 | 112 | 7 | 87 | 10 | 199 (8) |
| \$40,000 - \$49,999 | 71 | 5 | 42 | 5 | 113 (5) |
| \$50,000 - \$59,999 | 44 | 3 | 24 | 3 | 68 (3) |
| \$60,000 - \$69,999 | 30 | 2 | 20 | 2 | 50 (2) |
| ≥ \$70,000 | 53 | 4 | 25 | 3 | 78 (3) |
| **Regularly bitten by mosquitos** | 1463 | 98 | 824 | 98 | 2304 (98) |
| **Female** | 1039 | 69 | 547 | 65 | 1596 (68) |
| **Exposure to educational campaigns, last 12 months** | 368 | 25 | 219 | 26 | 587 (25) |
|  | **Mean** | **SD** | **Mean** | **SD** | **Mean (SD)** |
| **Age** | 37 | 8.8 | 37 | 8.8 | 37 (8.8) |
| **Household Expenditure on Mosquito Products** | \$151 | \$195 | \$244 | \$259 | \$185 (\$225) |

<sup>a</sup> There were 6 household-level mosquito-preventive behaviors total: (1) Eliminating stagnant water around the house (ex: cleaning containers that collect water like flowerpots and/or gutters; (2) Covering containers that collect water; (3) Emptying trash cans, drums, or in-ground trash cans if they had standing water; (4) Cleaning or removing tires and/or debris from yard; (5) Spraying insecticide or fumigate indoors or outdoors (ex: Blackjack, Real Kill, Raid); (6) Burning citronella candles and/or mosquito coils.
<sup>b</sup> Percentages presented as proportion of non-missing observations. Total number of missing observations on each variable are as follows: a) Number of preventive behaviors engaged in (outcome variable), 17; b) education level, 33; c) mosquito bite frequency, 4; d) sex, 0; e) income level, 159; f) exposure to educational campaigns, 35; g) age, 0; h) household expenditure on products, 3.

The regression models that encoded risk perception as an ordinal exposure variable (Models 1, 3, 5, and 7) produced similar outputs to those in which the exposure was coded as disjoint indicator variables (Models 2, 4, 6, and 8). Nonetheless, the Bayesian information criterion (BIC) and Akaike information criterion (AIC) values (see S1 Table) preferred the models in which the exposure is coded as an ordinal variable (Models 1, 3, 5, and 7). These models also produced estimates with greater precision (Figures 1 & 2, S1 Table). Therefore, the subsequent description of results will focus on reporting model outputs from models that used ordinal coding of the exposure variable and multiple imputation (models 3 and 7).

**Figure 1.**
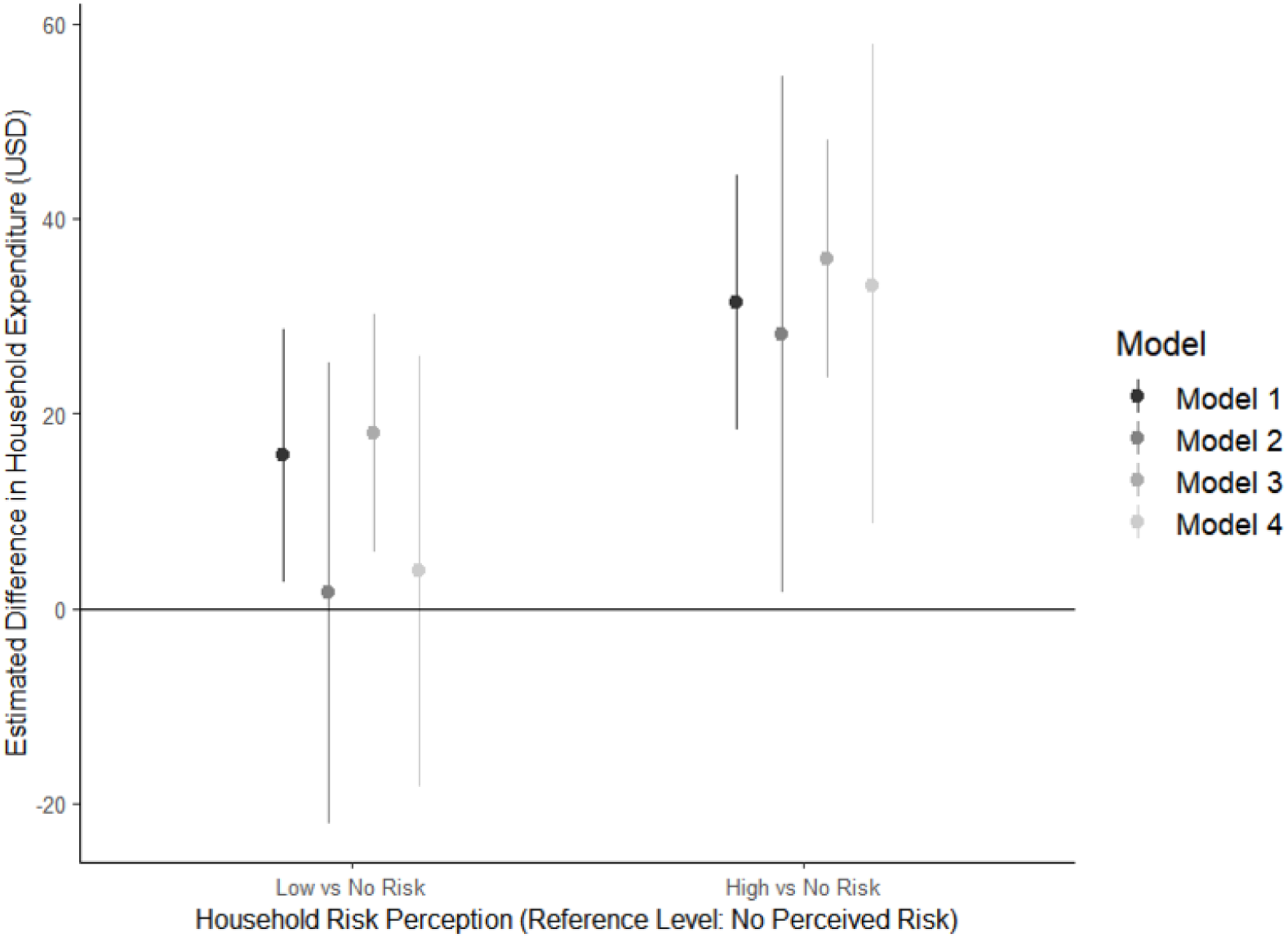
Estimates and 95% confidence intervals from four models of the difference in household expenditure (USD) on mosquito-preventive products per risk perception group, COPA, 2018-2019^a^ ^a^ All models included the full set of covariates as described in the methods section. Point estimates (dots) represent the average difference in household expenditure (USD) on mosquito preventive products when the household representative perceived their household at either low risk or high risk (based on the x-axis) compared to when the representative perceived their household at no risk. The bands around each dot represents the 95% confidence intervals for each point estimate. Positive values indicate increased spending; negative values indicate decreased spending. Model 1 used an ordinal exposure variable and complete case analysis; Model 2 used disjoint indicator coding for the exposure variable and complete case analysis; Model 3 used ordinal exposure variable and multiple imputation to address missing observations; Model 4 used disjoint indicator coding for the exposure variable and multiple imputation. Model fit statistics can be found in Table 4 in the supplementary material. Based on superior model fit statistics, Model 3 results are reported in the text.

Figure 1 shows the output from the regression analysis for the first outcome of interest, household expenditure on mosquito-preventive products and services. The adjusted linear regression model with ordinal exposure coding (Model 3) demonstrated that a 1-unit increase in dengue risk perception (from no risk perception to low, or from low to high risk perception) was associated with a $17.9 (95% CI: $5.8, $30.1) increase in annual household expenditure to prevent mosquito bites. On average, households where the household representative perceived their household at high risk of contracting dengue in the next year spent $35.9 ($23.7, $48.1) more annually on mosquito bite prevention compared to a household where the household representative considered their household at no risk. Figure 1 provides a graphical representation of the model estimates and 95% confidence intervals. For a detailed comparison of Model 1 – Model 4 outputs, see S1 Table.

**Table 4.**
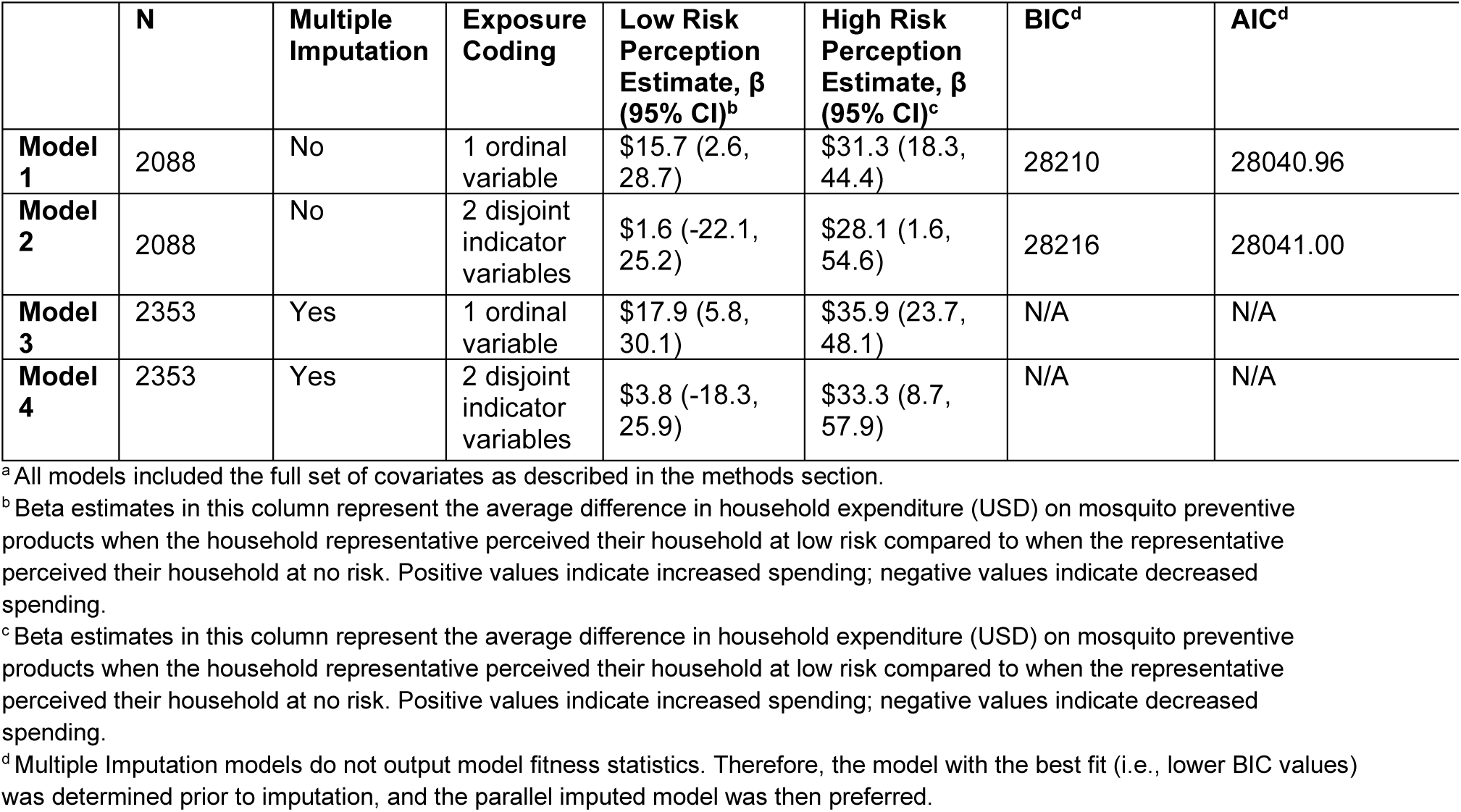
Model outputs describing the relationship between perceived risk and household expenditure (USD) on mosquito-preventive products, COPA, 2018-2019^a^.

The regression model results for the second outcome of interest—the probability of engaging in at least three household-level protective behaviors—are shown in Figure 2. This second set of regression models demonstrated that the probability of engaging in at least three household-level mosquito-preventive behaviors also increased with increasing arboviral disease risk perception. Model 7, which used ordinal exposure coding and multiple imputation, estimated that each 1-unit increase of risk perception was associated with an absolute increase of 5.1 percentage points (95% CI: 0.02, 0.08) in the probability that the household would report engagement in three or more household-level mosquito-preventive behaviors. Thus, households with a household representative who reported a high risk perception had a probability of engaging in three or more household-level mosquito-preventive behaviors that was 10.2 percentage points greater (95% CI: 7.2, 13.0), compared to households in which the representatives perceived their household at no risk of arboviral disease. Figure 2 provides a graphical representation of the model estimates and 95% confidence intervals. For a detailed comparison of Model 5 – Model 8 outputs, see S1 Table.

**Figure 2.**
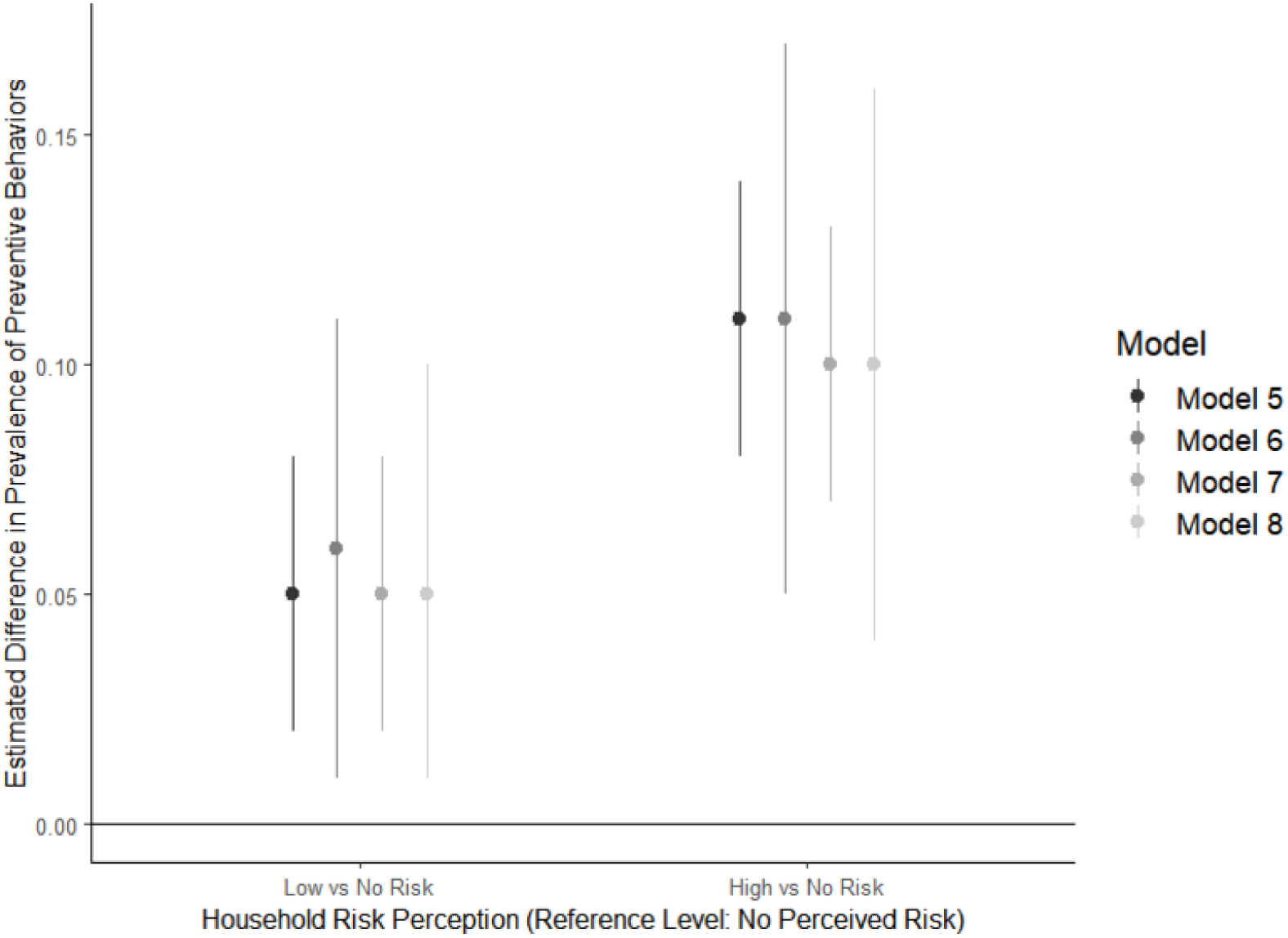
Prevalence difference estimates and 95% confidence intervals from four models representing the probability of household engagement in at least three protective behaviors per risk perception group, COPA, 2018-2019^a^ ^a^ All models included the full set of covariates as described in the methods section. Point estimates (dots) represent the average difference in probability that the household engaged in 3 or more protective behaviors when the household representative perceived their household at either low risk or high risk (based on the x-axis) compared to when the representative perceived their household at no risk. The bands around each dot represents the 95% confidence intervals for each point estimate. Positive values indicate increased spending; negative values indicate decreased spending. Model 5 used an ordinal exposure variable and complete case analysis; Model 6 used disjoint indicator coding for the exposure variable and complete case analysis; Model 7 used ordinal exposure variable and multiple imputation to address missing observations; Model 8 used disjoint indicator coding for the exposure variable and multiple imputation. Model fit statistics can be found in Table 5 in the supplementary material. Based on superior model fit statistics, Model 7 results are reported in the text.

## Discussion

This study analyzed a large (N=2,353) sample of households in Ponce, Puerto Rico, with a median annual household income of $10,000-$19,999. This analysis found that after adjusting for income and other confounders, households with high perceived risk of contracting arboviruses reported spending $35.9 more per year (95% CI: $23.7, $48.1) on preventive products and services compared to households with no perceived risk. In a population where the median annual household income was less than $20,000, these results demonstrate that households are willing to invest in mosquito-bite prevention despite potentially limited resources if there is a high perceived risk of arboviral disease. Study data also showed a positive association between the household representative’s perception of risk and the household’s probability of engaging in at least three risk-reducing behaviors. These findings highlight the role of arboviral risk perception in influencing risk-reducing behaviors and support the continued development of community-based programs to increase awareness of arboviral disease risk across Puerto Rico.

**Table 5.** Model outputs describing the relationship between perceived arboviral risk and the probability of engaging in at least three household-level protective behaviors^a^. COPA, 2018-2019.

| | N | Multiple Imputation | Exposure Coding | Low Risk Perception Estimate, $\beta$ (95% CI)<sup>b</sup> | High Risk Perception Estimate, $\beta$ (95% CI)<sup>c</sup> | BIC<sup>d</sup> | AIC<sup>d</sup> |
| --- | --- | --- | --- | --- | --- | --- | --- |
| **Model 5** | 2093 | No | 1 ordinal variable | 0.05 (0.02, 0.08) | 0.11 (0.08, 0.14) | 2831.1 | 2661.7 |
| **Model 6** | 2093 | No | 2 disjoint indicator variables | 0.06 (0.01, 0.11) | 0.11 (0.05, 0.17) | 2838.6 | 2663.5 |
| **Model 7** | 2353 | Yes | 1 ordinal variable | 0.05 (0.02, 0.08) | 0.10 (0.07, 0.13) | N/A | N/A |
| **Model 8** | 2353 | Yes | 2 disjoint indicator variables | 0.05 (0.01, 0.10) | 0.10 (0.04, 0.16) | N/A | N/A |

<sup>a</sup> All models included the full set of covariates as described in the methods section.
<sup>b</sup> Beta estimates in this column represent the average difference in probability that the household engaged in 3 or more protective behaviors when the household representative perceived their household at low risk compared to when the representative perceived their household at no risk. Positive values indicate increased probability of engaging in household-level protective behaviors; negative values indicate decreased probability of engaging in household-level protective behaviors.
<sup>c</sup> Beta estimates in this column represent the average difference in probability that the household engaged in 3 or more protective behaviors when the household representative perceived their household at high risk compared to when the representative perceived their household at no risk. Positive values indicate increased probability of engaging in household-level protective behaviors; negative values indicate decreased probability of engaging in household-level protective behaviors.
<sup>d</sup> Multiple Imputation models do not output model fitness statistics. Therefore, the model with the best fit (i.e., lower BIC values) was determined prior to imputation, and the parallel imputed model was then preferred.

The association between heightened arbovirus risk perception and increased prevalence of behaviors to prevent arboviral infections in this study fits well with previous research related to community engagement in mosquito vector control in Puerto Rico. For example, in a CDC case study of dengue control measures during an epidemic in the 1990s, visible public spraying efforts, while largely ineffective, reduced residents’ engagement in mosquito bite preventive activities [21]. As residents shifted the responsibility of mosquito control to the government, they perceived themselves at low risk of arboviral infection and were less likely to engage in further protective behaviors. Moreover, a pilot study conducted in Manuel A. Pérez Public Housing in San Juan, Puerto Rico in 2018 demonstrated the potency of community-based participatory campaigns to increase employment of mosquito-control tactics [22]. In that study, community members were directly engaged in the process of planning and implementing a risk communication initiative for their community. After the initiative, there was a significant increase in community members’ recognition of personal and community responsibility for the prevention of mosquito-borne disease, and their engagement in preventive behaviors for mosquito control increased [22].

Key strengths of this study include its sample size and the detailed information captured for each household, which allowed for comprehensive analyses. Another strength of this analysis is the use of multiple imputation, which allowed us to use all 2,353 households in our analysis and mitigate the risk of selection bias introduced by complete case analyses [46,47].

As in most studies on attitudes and practices that utilize in-person interviews, there may be some desirability bias in exposure and outcome measurement. Specifically, because the data collection teams visited participants at home after enrolling in a study focused on arboviruses, participants may have overreported protective behaviors, particularly if they felt that these were actions expected or supported by study staff. Likewise, participants who truly perceived themselves at no risk may have reported a low perceived risk of dengue to satisfy perceived interviewer expectations.

Another limitation of our study is the cross-sectional nature of the data, which does not allow us to assess the temporal sequence between the exposure and outcomes. While this study assessed whether households perceiving themselves at high risk would participate in preventive behaviors, it is possible that some participants perceived themselves at low or no risk of arbovirus in the upcoming 12 months because of their high engagement in prevention in the previous 12 months. Nevertheless, if this were the overarching trend, we would have expected to see a negative correlation between the exposure and outcome variables named in this analysis [27]. One final limitation in this study is residual confounding bias in the mosquito bite variable. Due to data unavailability, we dichotomized this variable, but there is plausible heterogeneity within the “bitten” category, particularly when compared to the “never bitten” level.

Our findings add to the growing body of evidence in support of community education interventions for populations at risk of contracting arboviral diseases. Paired with other findings [19,21,22], our results suggest that key stakeholders should invest in community-based participatory approaches to mosquito control, with particular focus on providing information in an accessible manner for community members to accurately interpret their risk and make more informed choices to reduce their risk of arboviral disease.

## Data Availability

Due to data security and confidentiality guidelines, all analyses of COPA data must be formally requested to CDC and PHSU. External researchers can request access to a restricted use dataset after submitting a concept proposal. Data requests related to COPA can be sent to.

